# Spatiotemporal Response Heterogeneity Across Metastatic Lesions Informs Drug Efficacy and Patient Survival in Colorectal Cancer

**DOI:** 10.1101/2020.10.19.20215160

**Authors:** Jiawei Zhou, Quefeng Li, Yanguang Cao

## Abstract

The sum of target lesions is routinely used to evaluate patient objective responses to treatment in the RECIST criteria, but it neglects the response heterogeneity across metastases. This study argues that the spatiotemporal response heterogeneity across metastases informs drug efficacy and patient survival. We analyzed the longitudinal data of 11,404 metastatic lesions in 2,802 colorectal cancer patients and examined their response heterogeneity. The response dynamics of metastatic lesions varied broadly across anatomical locations and therapies. High inter-lesion heterogeneity is associated with worse survival (p < 0.001), while targeted therapies (bevacizumab or panitumumab) reduced the inter-lesion heterogeneity (p < 0.05) and elicited more favorable effects on liver lesions (p < 0.001) than chemotherapy alone. The responses of liver lesions predicted patient survival more significantly than the lesions in the lungs and lymph nodes. Altogether, the high spatiotemporal heterogeneity across metastases should be integrated into current methods for treatment evaluation and patient prognosis.

**Significance:** The spatiotemporal heterogeneity across metastases in response to first-line therapies in colorectal cancer is informative for drug efficacy and patient survival, particularly in targeted therapy. Our findings provide evidence to support the inclusion of individual lesion response in the RECIST to improve the assessment of drug efficacy and patient survival.

## Introduction

The systemic spread of cancer cells from the primary site to distant organs is the leading cause of cancer modality, accounting for about 90% of cancer deaths (1). Preventing metastasis remains the main but challenging objective in cancer therapy (2). Although originating from the same primary tumor, metastatic lesions at different anatomical sites usually possess different tumor characteristics (3,4), due to divergent somatic mutations, epigenetic alterations, and organ-specific microenvironments (5,6). The genomic and phylogenetic heterogeneity across metastatic lesions have been illustrated in-depth using genomic sequencing data coupled with advanced computational approaches (7,8). However, the phenotypic heterogeneity in response to interventions across metastases and its correlation with patient prognosis has not been well characterized, which is the focus in our study.

The magnitude of therapeutic efficacy is routinely quantified based on overall tumor burden, calculated by the sum of all target lesions according to the Response Evaluation Criteria in Solid Tumors (RECIST) v1.1 (9). The RECIST guidelines, adopted in most clinical trials, do not account for the response disparities across metastatic lesions and may not be optimal for targeted therapies (10). Unlike conventional cytotoxic therapy, the responses of targeted therapies and acquired resistance are often lesion-specific. In a study of non-small cell lung cancers, the benefits of nivolumab varied broadly across metastatic sites. The efficacy was more pronounced in the lymph nodes (LN) lesions than in the liver and bone lesions (11). In another study, the patterns of response and progression displayed high lesion-specific variation in *BRAF*-mutant metastatic melanoma patients treated with a combination of two MAPK targeted therapies, dabrafenib and trametinib (12). Response variations across anatomical sites could result from divergent clonal genetic, epigenetic, and transcriptional features (13), conferred by different evolutionary pressures and organ-specific tumor microenvironments (14). Using the sum of target lesions may conceal the unique phenotypes of each lesion and the lesion-selective effect of targeted therapy, thus fail to disclose insights into lesion-specific response and resistance. We argue that the quantitative assessment of the spatiotemporal response across metastatic lesions can add another dimension to tumor heterogeneity and resistance, thereby deepening the investigation of the organ-specific factors that shape the phenotypes of metastases.

In our analysis, we considered colorectal cancer (CRC) as the tumor system. CRC has a high prevalence and mortality and is expected to cause approximately 53,200 deaths in 2020 (15). The five-year survival rate is only 13.1% for metastatic colorectal cancer (mCRC) in contrast to 90.1% for localized CRC patients (16). Many advances have been made in CRC treatment, with the use of targeted therapies combined with the standard chemotherapies. Targeted therapies, including anti-EGFR and anti-VEGF antibodies and recently approved anti-PD-1/PD-L1 immunotherapies, provided a durable response and prolonged patient survival in a selected group of patients (17). This study incorporated longitudinal data from five Phase III trials, comprising 2,802 mCRC patients with 11,404 metastatic lesions. We examined the spatiotemporal response heterogeneity across metastases in either chemotherapy alone or combined with targeted therapies. We corroborated the associations between lesion-specific response and patient progression-free survival (PFS), overall survival (OS), and *KRAS* status. Our findings supported the substantial significance of including spatiotemporal response dynamics across metastases for the improved evaluation of drug efficacy and patient prognosis.

## Results

### The response dynamics were heterogeneous across metastatic lesions

We evaluated the response dynamics of all individual metastatic lesions (n =11,404), as well as the total tumor burden (the sum of the diameter of the target lesions within each patient) from 2,802 patients for comparison (Fig. 1A). Patient demographics and clinical characteristics are summarized in Table 1. We defined three responsive phases (response, stable, relapse) according to the RECIST v1.1 and developed an algorithm to quantify the durations of these phases (see Methods and Supplementary Fig. S1). The responsive phases, together with the tumor nadir size (best response) and the last tumor size before the end of the trial (relapse rate), were used to assess the lesion-level response to therapy (Fig. 1B). As shown in Fig. 1C, the dynamics of the total tumor burden masked the individual lesion responses. For instance, in patient 508, the total tumor burden exhibited an initial response, then a stable response, and finally, the relapse phase, while the LN metastatic lesion remained in the stable phase throughout the treatment. In patient 501, the total tumor burden remained stable throughout the trial, while the abdomen lesion quickly relapsed, and the gastrointestinal (GI) tract lesion responded durably during the treatment. The response dynamics could also be different for lesions within the same organ, as shown in patient 1312. The patients whose lesions responded differently from the total tumor burden are summarized in Fig. 1D. More than 60% of the patients had at least one lesion responding differently from the total tumor burden. In addition, the fraction of lesions with differently responding dynamics varied across the anatomical sites, with the lowest fraction in the liver (35%) and the highest in the bone (48%) (Fig. 1E). The small fraction of differently responding lesions in the liver was mainly due to the large size and number of hepatic metastases dictating the trend of the total tumor burden. Collectively, these observations suggested that the response status of the total tumor burden could not fully reflect the response dynamics of the individual lesions, as it failed to account for the lesion-specific response and resistance features.

**Table 1.** Patient demographic information.

| Demographics |  | Total (n = 2,802) |
| --- | --- | --- |
| Male, n (%) |  | 1,688 (60.2) |
| Age (years), mean (sd) |  | 60.74 (10.91) |
| White or caucasians, n (%) |  | 2,611 (93.2) |
| BSA, mean (sd) |  | 1.87 (0.24) |
| LDH baseline $\geq 1.5$ UNL, n (%) | | 836 (29.8) |
| ECOG performance, n (%) |  |  |
|  | 0 | 1,536 (54.8) |
|  | 1 | 1,178 (42.0) |
|  | 2 | 87 (3.2) |
| *KRAS*, n (%) |  |  |
|  | Wild-type | 795 (28.4) |
|  | Mutant | 593 (21.2) |
|  | Unknown | 1,414 (50.4) |
| Primary tumor Type, n (%) |  |  |
|  | Colon | 1910 (68.2) |
|  | Rectum | 892 (31.8) |
| Number of metastatic sites, n (%) |  |  |
|  | 1 | 382 (13.6) |
|  | 2 | 449 (16.0) |
| | $\geq 3$ | 1971 (70.4) |
| Liver-metastases only, n (%) |  | 1347 (48.1) |
| Prior surgery, n (%) |  | 2535 (90.5) |
| Prior radiation therapy, n (%) |  | 367 (13.1) |
| Previous adjuvant therapy, n (%) |  | 505 (18.0) |
| RECIST response, n (%) |  |  |
|  | Complete response | 77 (2.7) |
|  | Partial response | 1,069 (38.2) |
|  | Stable disease | 1,173 (41.9) |
|  | Progressive disease | 401 (14.3) |
|  | Not detected | 31 (1.2) |
|  | Unable evaluate | 39 (1.4) |
|  | Unknown | 12 (0.3) |
| Treatment, n (%) |  |  |
|  | Bevacizumab plus chemotherapy | 377 (13.5) |
|  | Bevacizumab plus panitumumab plus chemotherapy | 371 (13.2) |
|  | FOLFIRI | 419 (15.0) |
|  | Panitumumab plus FOLFIRI | 425 (15.2) |
|  | FOLFOX | 769 (27.4) |
|  | Panitumumab plus FOLFOX | 441 (15.7) |

**Figure 1.**
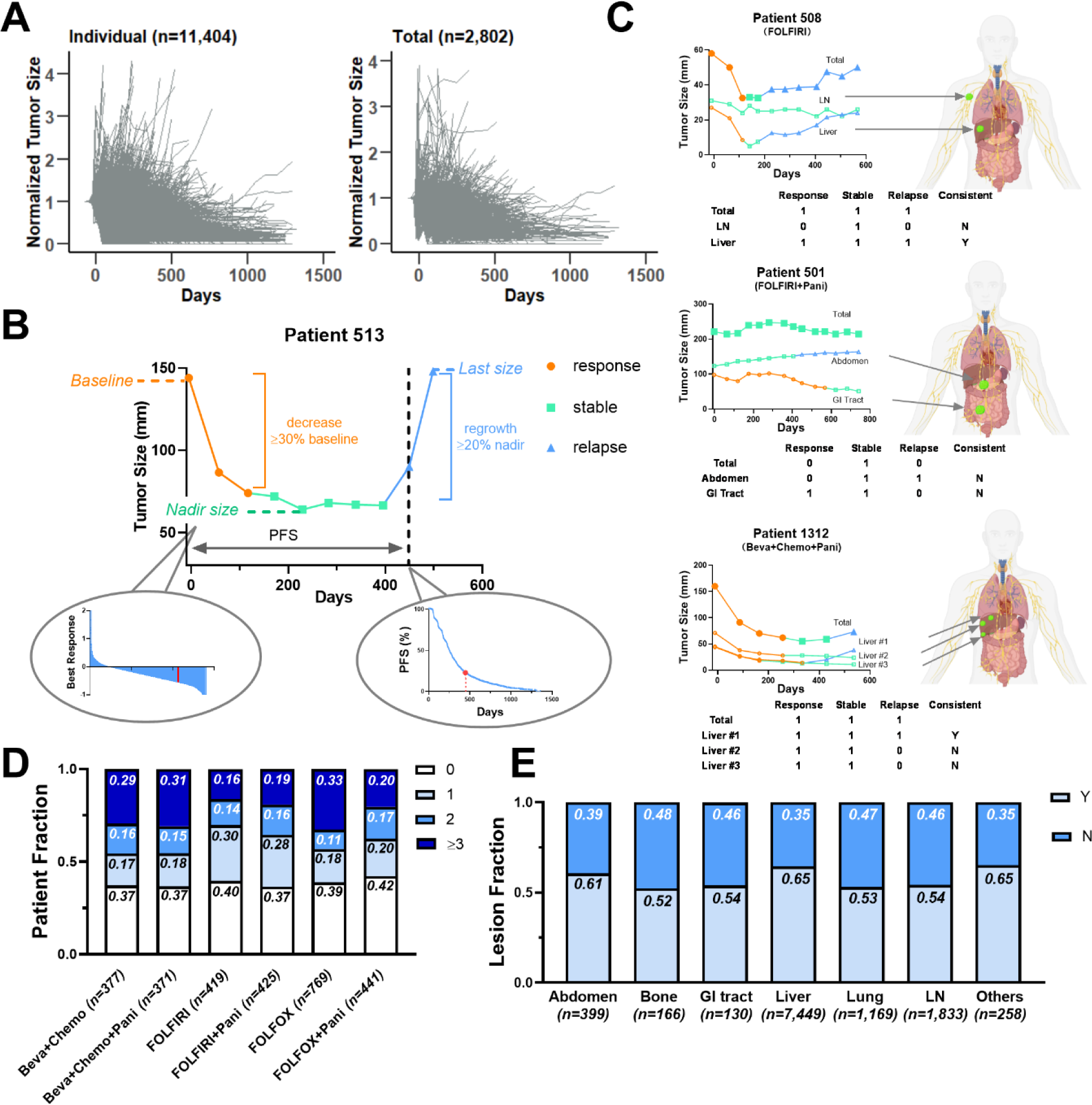
Heterogeneous response dynamics across metastatic lesions. (**A**) Spider plots of individual tumor size (left) and total tumor burden (right) normalized by tumor baseline during the course of treatment. (**B**) Five variables (response, stable, and relapse phase, tumor nadir size, and last size before the end of trials normalized by the baseline) were extracted to describe tumor response patterns. (**C**) Total tumor burden and individual metastatic lesion response dynamics during the treatment in three representative patients, patient 508, 501, and 1312. The lesion response, stable and relapse phases are labeled in orange, green and blue respectively. Three binary variables (Response, Stable, Relapse) represent whether their corresponding phases existed (= 1) or not (= 0). Lesions were defined as inconsistent (N) if their phase variables were not consistent with those in the total tumor burden. Otherwise, they would be defined as consistent (Y). (**D**) The fractions of patients with 0, 1, 2, or ≥ 3 inconsistent metastatic lesions across treatments. (**E**) The fractions of lesions inconsistent (N) or consistent (Y) with the total tumor burden across the anatomical sites.

### The heterogeneous responses across metastatic lesions predicted patient survival

To quantify response heterogeneity, we computed the Gower distance (18), an index reflecting the degree of divergence in the response dynamics across metastases. Five parameters extracted from the lesion response profiles were used to calculate the lesion-level Gower distance, including three phases (response, stable, and relapse), tumor nadir ratio, and last ratio (see Methods and Fig. 2A). Patient-level Gower distance was derived by averaging the lesion-level Gower distances for all metastases within each patient, and it was independent of the lesion number for patients with more than two lesions (p = 0.43 in ANOVA). The lesion response dynamics of the patients with high or low Gower distances are shown in Supplementary Fig. S2, where a higher Gower distance implies a higher inter-lesion heterogeneity (ILH) in the response dynamics.

**Figure 2.**
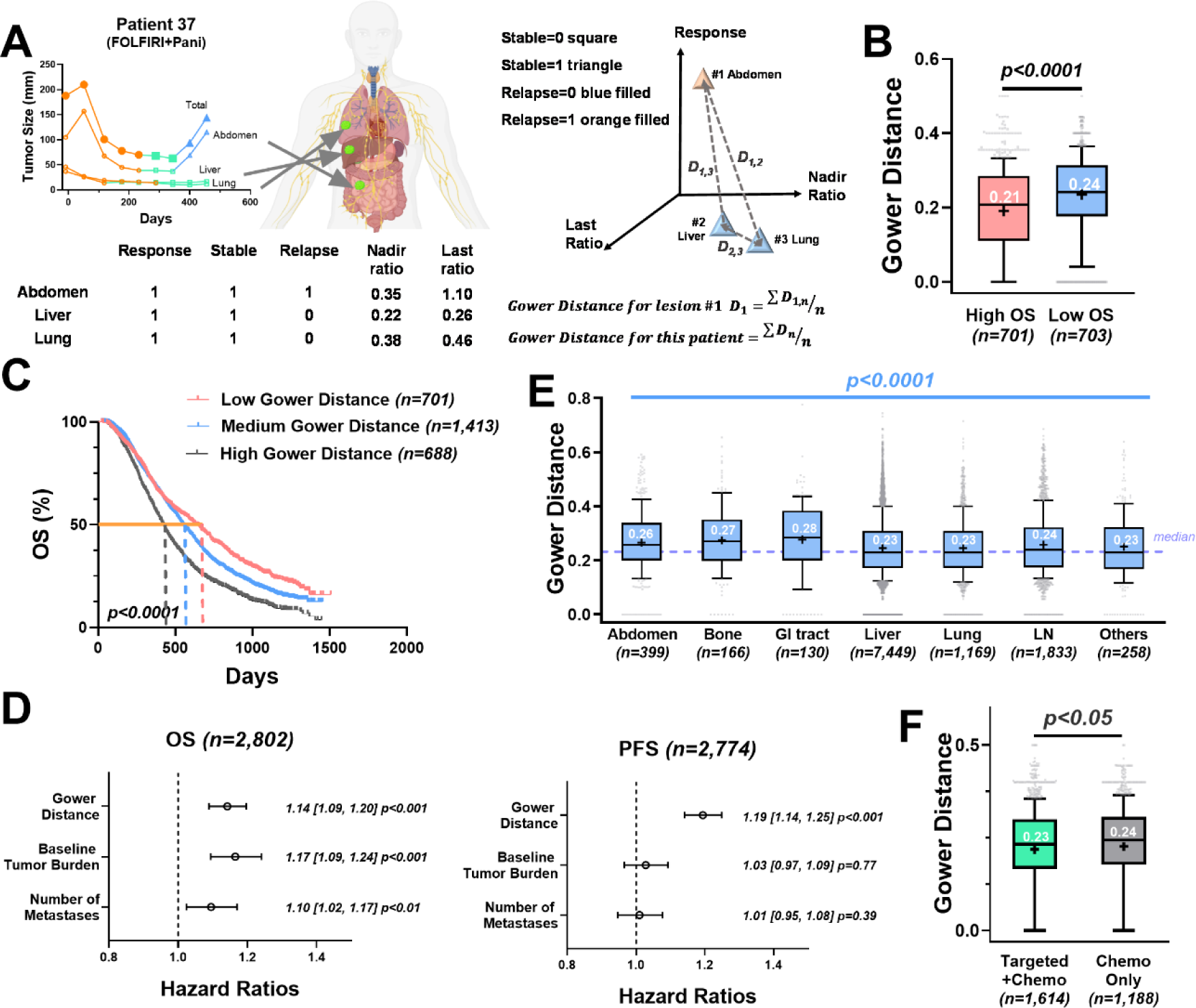
Response heterogeneity across metastatic lesions is associated with survival. (**A**) Five parameters extracted from the lesion response patterns, including three phase variables (response, stable, and relapse), nadir ratio (baseline-normalized nadir), and last ratio (baseline-normalized last size), were used to calculate the Gower distance. (**B**) Boxplot of the Gower distances in patients with top 25% overall survival (high OS) versus patients with bottom 25% overall survival (low OS). (**C**) Kaplan-Meier curves of patient OS grouped by Gower distance (25^th^ and 75^th^ quantiles as the cut-offs). (**D**) Forest plot of the Cox proportional hazards model stratified by treatment type on the OS and PFS, controlling for baseline tumor burden and number of metastases. The hazard ratio with a 95% confidence interval and the p value of each covariate is labeled in the plot. (**E**) Boxplot of lesion-level Gower distances grouped by anatomical site. The dashed purple line represents the median of the Gower distances. (**F**) Boxplot of patient-level Gower distances in targeted therapy combined with chemotherapy (Targeted + Chemo) versus chemotherapy alone (Chemo). (B, E, F) The box extends from the 25^th^ to 75^th^ percentiles and the line in the middle is plotted as the median (labeled in white). The mean of the group is plotted as the symbol “+”. The whiskers are drawn down to the 10^th^ percentile and up to the 90^th^ percentile. Points below and above the whiskers are drawn as gray individual points.

To examine the association between patient survival and ILH in the response dynamics, we stratified patients by their OS. The patients with higher OS had lower ILH in their response dynamics compared to those with relatively lower OS (p < 0.0001, Fig. 2B). Similarly, the patients with high ILH had significantly worse OS (median 422 days) than those with medium ILH (OS 550 days) or low ILH (OS 630 days) (p < 0.0001, Fig. 2C). This observation was confirmed in the Cox proportional hazard model stratified by therapy (Gower distance HR=1.14, p < 0.001 in OS; HR=1.19, p < 0.001 in PFS), controlling the influence of the tumor baseline and the number of metastases (Fig. 2D). Overall, the increased ILH was associated with worse survival outcomes for patients with mCRC. These findings aligned with previous studies showing that tumors with higher response heterogeneity were associated with worse survival (19,20).

Next, we explored the spatial heterogeneity by comparing the lesion-level Gower distances across the metastatic organs. The organ with the highest Gower distance was the GI tract (median 0.28), and those with the lowest were the liver and the lungs (median 0.23) (Fig. 2E and Supplementary Table S2). The ILH also differed within the treatment types. Targeted therapies, including bevacizumab or panitumumab, into the standard chemotherapies, showed marked reductions in ILH (p < 0.05) compared to chemotherapy alone (Fig. 2F and Supplementary Table S3).

### Targeted therapies reduced lesion response heterogeneity and exhibited favorable efficacy on hepatic metastases

To further explore the response heterogeneity across treatments, we compared the ILH between the targeted therapies and chemotherapy alone. We analyzed the bevacizumab and the panitumumab arms to examine the lesion-level benefits conferred by the targeted therapies, as well as the resulting ILH. We analyzed 377 patients in the bevacizumab group, compared to the chemotherapy only group (769 patients in FOLFOX and 419 patients in FOLFIRI). As expected, the OS and PFS were significantly improved by bevacizumab (p < 0.0001), which was major evidence supporting bevacizumab’s regulatory approval (Fig. 3A). Patients with relatively lower Gower distances had significantly longer survival in the bevacizumab arm, reinforcing that a higher ILH is associated with worse survival (Fig. 3B). Next, we tested whether the survival benefit conferred by bevacizumab was associated with a reduction in lesion response heterogeneity. Interestingly, despite notably lower lesion nadir and last ratios and more extended response and stable durations (p < 0.0001, Supplementary Fig. S3 and Table S4), the Gower distances in the bevacizumab group were not significantly different from the chemotherapy group (p = 0.06, Fig. 3C and Supplementary Table S5). This was mainly due to high variability: the lesion-level Gower distances varied considerably across organs (Fig. 3D and Supplementary Table S6), which confounded the patient-level comparison of ILH, as shown in Fig. 3C.

**Figure 3.**
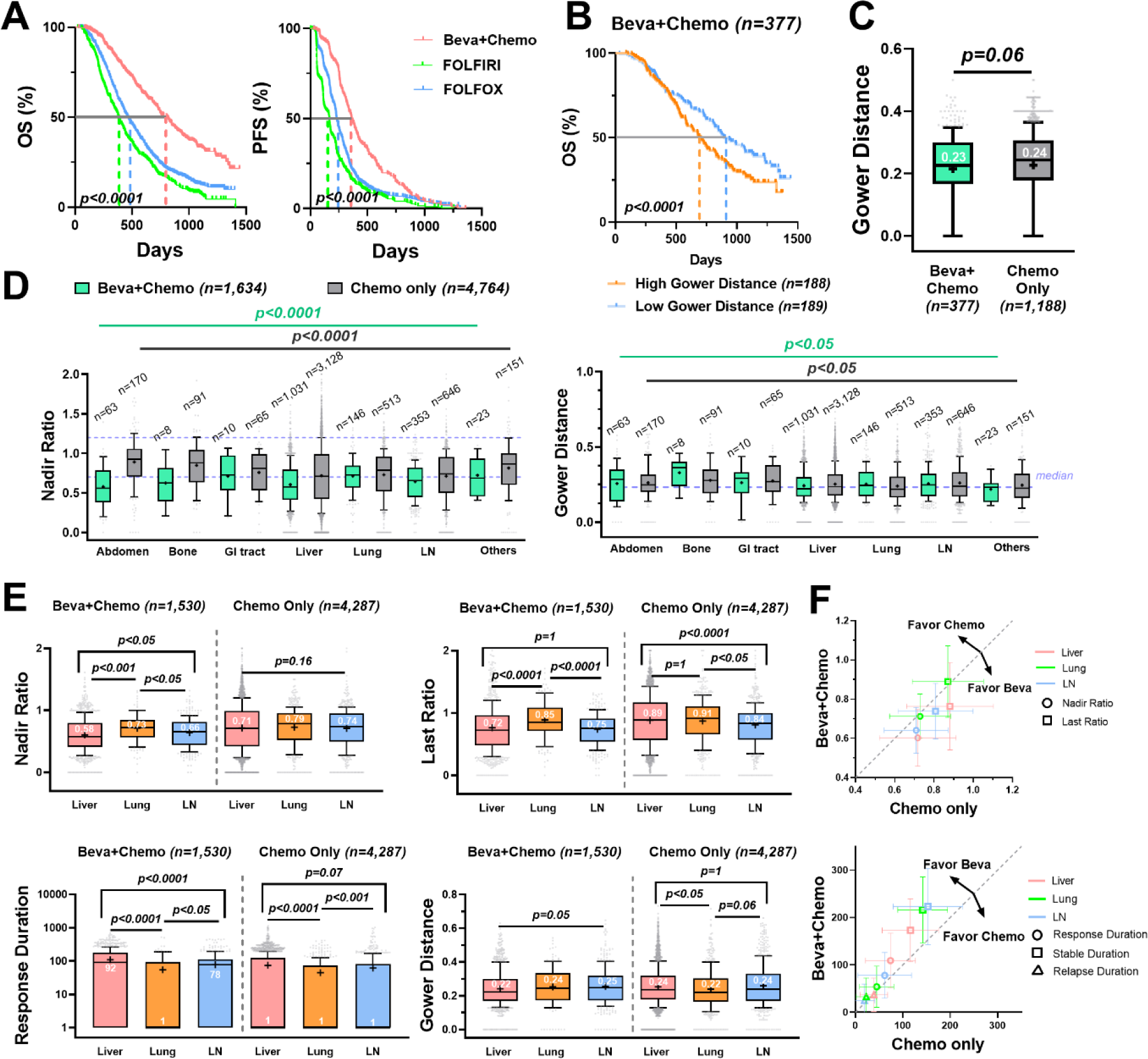
Bevacizumab reduced lesion response heterogeneity and exhibited favorable efficacy on hepatic metastases. Beva+Chemo: bevacizumab plus chemotherapy; Chemo: chemotherapy alone. (**A**) Kaplan-Meier curves of the OS (left) and PFS (right) in Beva + Chemo (n = 377), FOLFOLX (n = 769 in OS and n = 741 in PFS) and FOLFIRI (n = 419) group. (**B**) Kaplan-Meier curves of the OS grouped by Gower distances (median as the cut-off) in the Beva + Chemo group. (**C**) Boxplot of the patient-level Gower distances in Beva + Chemo versus Chemo (FOLFOX and FOLFIRI) group. (**D**) Boxplots of the lesions’ nadir ratios with the 0.7 and 1.2 values labeled in purple dashed line (left) and the lesions’ Gower distances with median labeled in purple dashed line (right) across anatomical sites. (**E**) Boxplots of the nadir ratio, last ratio, response duration, and Gower distance in liver (n = 1,031), lung (n = 146) and LN metastases (n = 353) in the Beva + Chemo (left) compared to the Chemo only (right, liver n = 3,128; lung n = 513; LN n = 646). The durations were all added with one to avoid zero values in logarithmic plots. (**F**) The mean ± 50% standard deviation of nadir ratio and last ratio (upper) and response, stable, and relapse durations (lower) in the Beva + Chemo versus the Chemo only group. The dashed gray line is y = x.

Next, we selected the top three metastatic organs for mCRC – liver, lung, and LN – for further analysis. Strikingly, in the bevacizumab arm, the liver lesions had a significantly better response, with lower nadir ratios, last ratios, and Gower distances, as well as a longer response duration than the lesions in the LN and lungs (Fig. 3E and Supplementary Table S7). In contrast, chemotherapy alone did not show noticeably better efficacy on the metastases in the liver than those in the LN and lungs. In the chemotherapy group, the liver lesions appeared to relapse faster than those in the LN, and showed higher heterogeneity than the lung metastases (Fig. 3E and Supplementary Table S8). As shown in Fig. 3F, compared to chemotherapy alone, the nadir and last ratios were much lower and the response durations were longer for all metastases in three major organs treated by bevacizumab. The highest efficacy on liver lesions was seen in the bevacizumab arm.

We performed a similar analysis of the panitumumab arm (866 patients), in comparison with the chemotherapy alone arm (FOLFOX or FOLFIRI). Significant benefits in both OS and PFS were observed when adding panitumumab to the chemotherapy (Fig. 4A). As in the bevacizumab group, a higher patient-level Gower distance was associated with worse patient survival in the panitumumab group (Fig. 4B). Panitumumab showed significantly better efficacy, compared to chemotherapy alone, with much lower tumor nadirs (p < 0.0005), last ratios (p < 0.01), and ILH (p < 0.05), as well as a more extended response phase (p < 0.0001) (Fig. 4C and Supplementary Fig. S4 and Table S9). The tumor nadir varied across organs, ranging from 0.64 (liver) to 0.92 (other) (Fig. 4D and Supplementary Table S10). The lesion response heterogeneity also varied across organs, which further differed between treatments (Fig. 4D and Supplementary Table S11). We further compared the response dynamics of the lesions in the three major organs. Similar to bevacizumab, panitumumab exhibited much lower nadir and last ratios and more extended response durations in the selected lesions than chemotherapy alone and had the best response in the liver metastases (Fig. 4E, Fig. 4F and Supplementary Table S12). Overall, a substantial response heterogeneity across anatomical lesions was observed and panitumumab showed remarkably better efficacy on liver metastases than chemotherapy alone.

**Figure 4.**
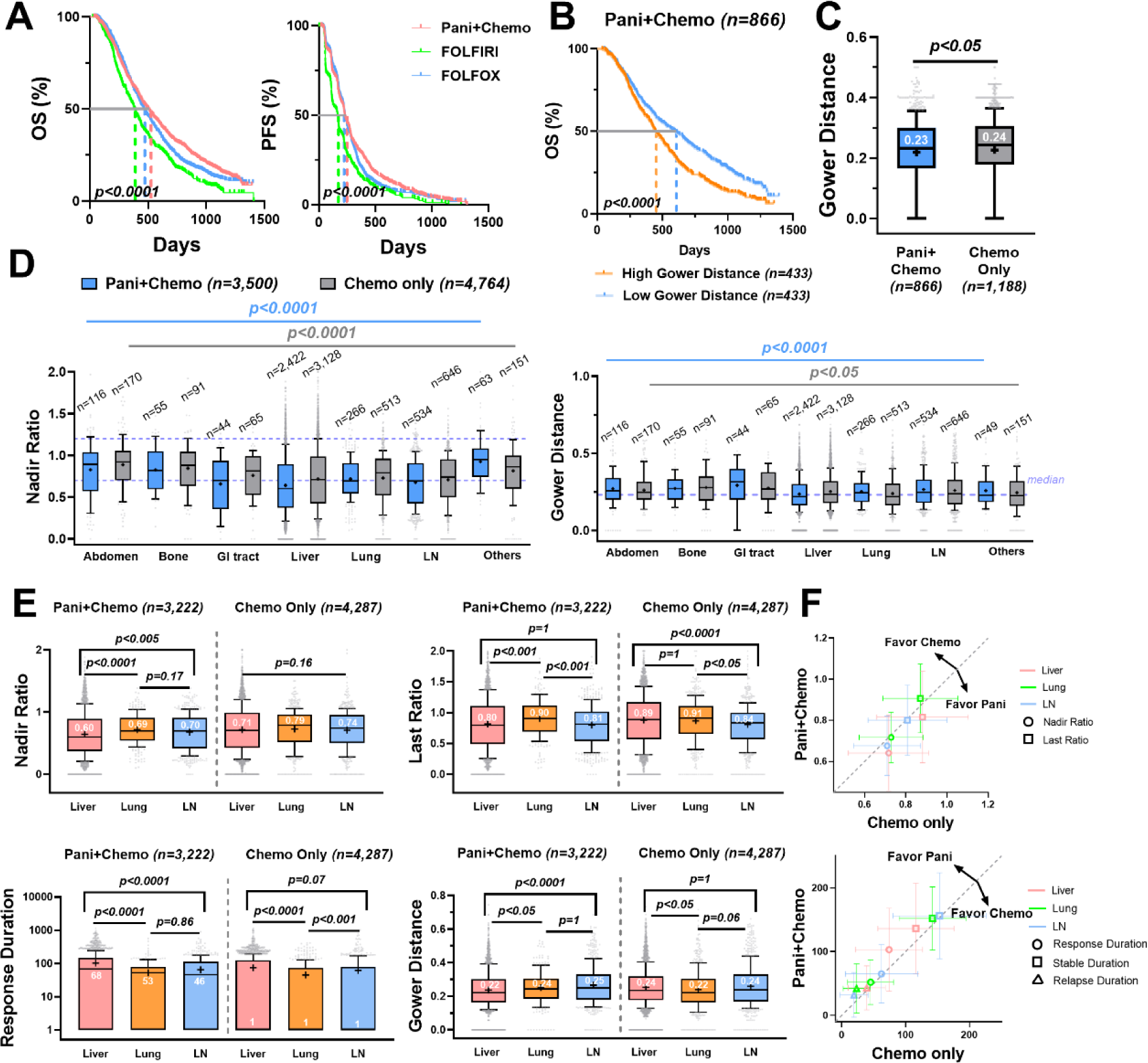
Panitumumab reduced lesion response heterogeneity and showed favorable efficacy on liver metastases. Pani + Chemo: panitumumab plus chemotherapy; Chemo: chemotherapy alone. (**A**) Kaplan-Meier curves of the OS (left) and the PFS (right) in the Pani + Chemo (n=866), FOLFOLX (n = 769 in OS, n = 741 in PFS) and FOLFIRI (n = 419) group. (**B**) Kaplan-Meier curves of the OS grouped by the Gower distances (median as the cut-off) in the Pani + Chemo group. (**C**) Boxplot of the patient-level Gower distances in Pani + Chemo versus Chemo (FOLFOX and FOLFIRI) group. (**D**) Boxplots of the lesions’ nadir ratios with the 0.7 and 1.2 values labeled in purple dashed line (left) and the lesions’ Gower distances with median labeled in purple dashed line (right) across anatomical sites. (**E**) Boxplots of the nadir ratio, last ratio, response duration, and Gower distance in liver (n = 2,422), lungs (n = 266) and LN metastases (n = 534) in the Pani + Chemo group (left) compared to the Chemo group (right, liver n = 3,128; lung n = 513; LN n = 646).(**F**) The mean ± 50% standard deviation of nadir ratio and last ratio (upper) and response, stable, and relapse durations (lower) in the Pani + Chemo versus Chemo only group. The dashed gray line is y = x.

### Lesion response was influenced by patient *KRAS* status

Anti-EGFR targeted therapies are prescribed for patients with wild-type *KRAS*. The interaction between *KRAS* status and lesion response was evaluated in this section. As expected, patients with *KRAS* mutation (n = 593) had significantly worse OS (p < 0.0001) and PFS (p < 0.01) compared to the wild-type patients (n = 765), which was consistent with previous findings that *KRAS* mutations in colon cancers have been associated with poorer survival and increased tumor aggressiveness (21). In the metastases of the three major organs, wild-type patients had relatively lower nadir and last ratios, and more extended response durations compared to the *KRAS* mutant patients (p < 0.0001, Supplementary Fig. S5). Since patients with *RAS* mutations were predicted to have a limited response to panitumumab (22), we further stratified the patients into either with or without panitumumab. The lesion-level comparisons showed that wild-type patients had significantly lower nadir and last ratios in hepatic metastases than their lung and LN metastases. In contrast, the liver-favorable effects were not evident in the *KRAS* mutant patients (Fig. 5A and Fig. 5B). Panitumumab significantly shrunk the liver and lung metastases, but to a lesser degree with the LN metastases. The efficacy of panitumumab was higher in the wild-type patients than in the *KRAS* mutated patients, mostly on hepatic metastases (Fig. 5C).

**Figure 5.**
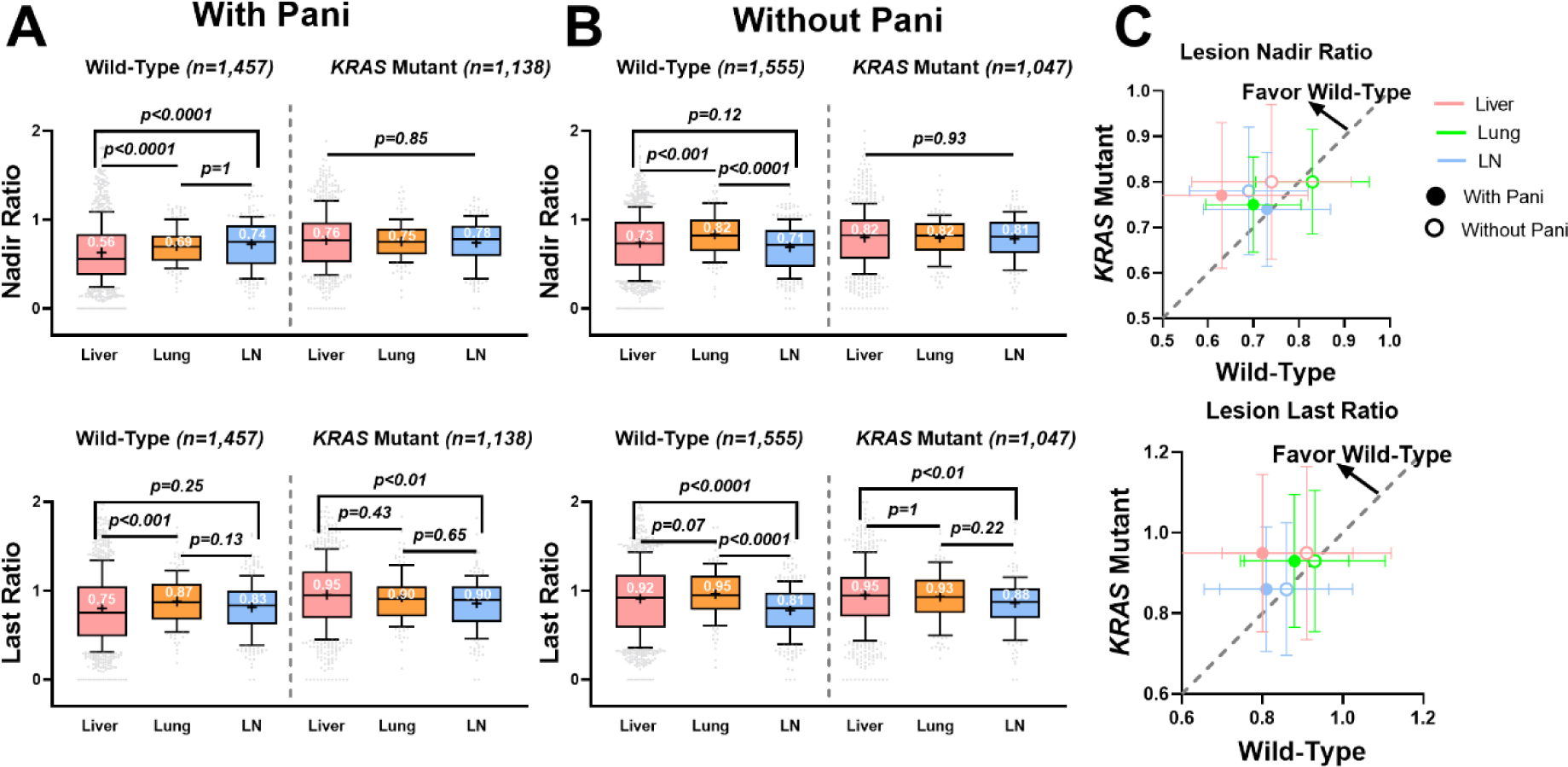
Lesion response heterogeneity was influenced by patient *KRAS* mutation status. (**A**) Boxplots of the nadir ratio (upper) and last ratio (lower) of the lesions in the wild-type versus *KRAS* mutant patients under panitumumab treatment (Beva + Chemo + Pani or FOLFIRI + Pani). (**B**) Boxplots of the nadir ratio (upper) and last ratio (lower) of the lesions in the wild-type versus *KRAS* mutant patients without panitumumab treatment (Beva + Chemo or FOLFIRI). (**C**) The mean ± 50% standard deviation of the nadir ratio (upper) and last ratio (lower) in the wild-type versus *KRAS* mutant patients. The dashed gray line is y = x.

### The response of hepatic metastases was closely associated with patient survival

To compare the survival benefit of organ-specific lesion response, we divided the patients with liver metastases (n = 2,229) into subgroups based on whether the shrinkage of the metastases in the liver or other organs was greater than 30% (the threshold for response in the RECIST v1.1). Patients with responding liver lesions (shrinkage > 30%) but non-responding lesions in other organs (shrinkage ≤ 30%) had significantly better OS (n = 142, median 546 days) and PFS (median 282 days) than those who had responding lesions in other organs but not in the liver (n = 80, OS 401 days; n = 79, PFS 176 days) (Fig. 6A).

**Figure 6.**
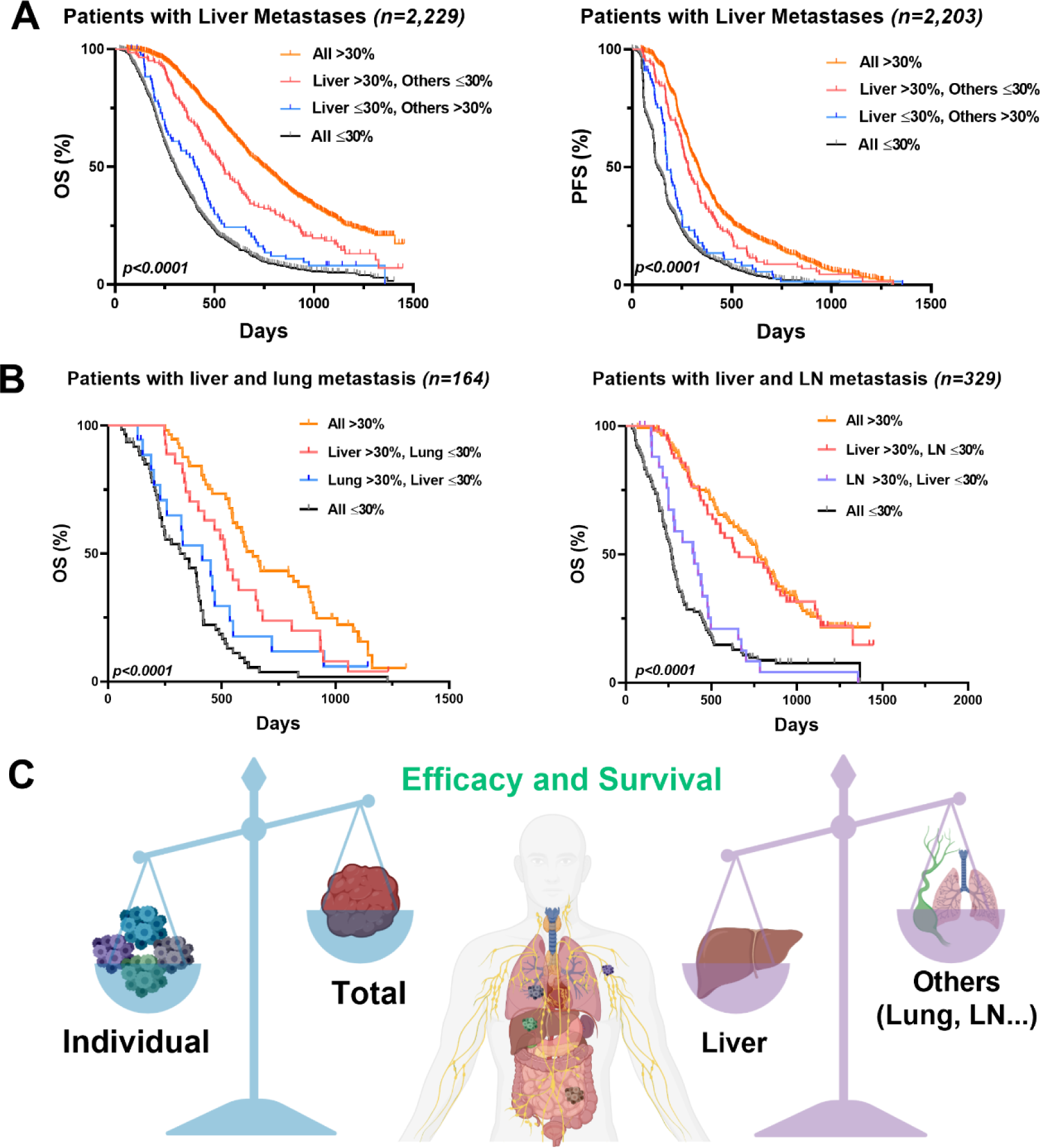
The response of liver lesions was more closely associated with patient survival. (**A**) Kaplan-Meier curves of the OS (left, n = 2,229) and the PFS (right, n = 2,203) in patients with liver metastases. The patients were divided into four sub-groups: all lesions shrinkage greater than 30% threshold (all > 30%, n = 1,113 in OS, n = 1,101 in PFS); liver lesions shrinkage > 30% while other lesions shrinkage ≤ 30% (liver > 30%, others ≤ 30%; n = 142); liver lesions shrinkage ≤ 30% while other lesions shrinkage > 30% (liver ≤ 30%, others >30%; n = 80 in OS, n = 79 in PFS); all lesions shrinkage ≤ 30% (all ≤ 30%, n = 894 in OS, n = 881 in PFS). (**B**) Kaplan-Meier curves of the OS in the patients with liver and lungs metastases only (left; n = 164), or the patients with liver and LN metastases only (right; n = 329). Similarly, patients were divided into four groups by the lesion shrinkage. In the left panel, all > 30% (n = 57); liver >30%; lung ≤ 30% (n = 27); lung >30%, liver ≤ 30% (n = 18); all ≤ 30% (n = 62). In the right panel, all > 30% (n = 138); liver >30%; LN ≤ 30% (n = 57); LN >30%, liver ≤ 30% (n = 27); all ≤ 30% (n = 107). (**C**) Due to the high inter-lesion heterogeneity, the response patterns of the total tumor sizes could not fully reflect the individual lesion response in the mCRC patients. The response dynamics of the liver lesions were more associated with survival than those of other organs in the mCRC patients.

We also analyzed the patient survival outcomes for those who only had liver and lung metastases (n = 164) or liver and LN metastases (n = 329). The patients with responding liver lesions but non-responding lung lesions had significantly extended OS (n = 27, median 519 days) than those with responding lung lesions but non-responding liver lesions (n = 18, OS 414 days). Similarly, the patients with responding liver lesions but non-responding LN lesions had longer OS (n = 57, median 662 days) than those with responding lung lesions but non-responding liver lesions (n = 27, OS 401 days) (Fig. 6B). In contrast, patients with responding LN or lung lesions but non-responding lesions in other organs had a much worse survival than those with other organ lesion shrinkage > 30% (Supplementary Fig. S6). These observations strongly suggested the different survival benefits of organ-specific lesion response. Altogether, patient survival was more closely associated with responding lesions in the liver than those in the LN and lungs (Fig. 6C).

## Discussion

Our study evaluated the spatiotemporal response heterogeneity (i.e., ILH) across 11,404 metastatic lesions in 2,802 mCRC patients. We characterized this substantial ILH and found that patients with higher ILH (i.e., high Gower distance) had worse survival outcomes. Metastases in different anatomical sites responded to the therapies differently. Metastatic lesions in the liver showed better and more uniform response to targeted therapies than lesions in the lungs and LN. Patients with *KRAS* mutations had less treatment benefits in their liver lesions than the wild-type patients. Regardless of therapy and *KRAS* status, a favorable response in the liver lesions was more associated with patient survival benefits than lesions at other sites. Our findings provided evidence of the importance of lesion-level response assessments and supporting the inclusion of lesion-level responses in the current RECIST approach for the improved evaluations of drug efficacy and patient survival outcomes in mCRC.

The lesion-level phenotypic response provided us another perspective to understand drug efficacy and resistance. These response differences across metastatic lesions might be attributed to the genetically and epigenetically divergent tumor cell clones within each lesion filtered by the organ-specific tumor immune microenvironment (23). Tumor development has been widely regarded as a process of Darwinian evolution (24). Many metastatic lesions, subject to different organ-specific evolutionary filters, have accumulated multiple genetically distinct sub-clones by the time of diagnosis (25). The clonal heterogeneity across metastatic lesions may result in different levels of sensitivity and resistance to treatments, leading to high phenotypic heterogeneity across metastatic lesions (26). The relatively uniform response in many long-term survivors could be partially due to similar genetic compositions, as well as reduced heterogeneity across metastatic lesions (27). Interestingly, the phenotypic diversity in lesions in the LN did not appear to be significantly higher than in the other distant lesions in our study, even though the genetic diversity was found to be much broader within LN metastases than distant lesions (8). *KRAS* mutation status showed a much higher impact on the response of liver lesions than lesions at the other sites. Although genotype-phenotype mapping was not within the scope of this study, our analyses reinforced the importance of the joint consideration of cancer genotyping with the lesion-level response phenotypes to better understand treatment efficacy and resistance. Overall, the current RECIST approach should be modified to extend beyond the sum of all target lesions, as it may potentially obscure the prognostic values of the unique response dynamics of individual metastases.

The favorable effect of targeted therapies on liver metastatic lesions could be ascribed to the high antibody penetrations in the liver compared to other organs. The liver is a highly perfused organ with sinusoidal vascular structures (28). Bevacizumab uptake in the liver and liver metastases was much higher than into the lungs and LN, according to an ^89^Zr-bevacizumab PET study (29). Similarly, panitumumab also exhibited a greater penetration into the liver than the lungs (30,31). Interestingly, the target expressions for two targeted therapies in liver metastases, VEGF and EGFR, were not higher than the LN and lung metastases in mCRC, indicating that the preferable uptake of antibodies in liver metastases was due to higher drug penetration but not enhanced target expression (32–34).

The liver is generally regarded as an immunologically tolerant organ, in which the hepatic adaptive immune cells become readily tolerogenic, promoting immunosuppressive tumor microenvironment (35). Therefore, the targeted therapies involving the activation of cytotoxic T cells (such as checkpoint blockades) are often less effective for liver metastases, compared to metastases in other anatomical sites (36,37). Unlike checkpoint blockades, the primary tumor-restraining mechanism of bevacizumab and panitumumab is to block or neutralize their respective antigens rather than eliciting effector functions (38). Different mechanisms of action helped explain why the targeted therapies evaluated in this study showed favorable effects on liver lesions compared to other organs. Intriguingly, the favorable benefit of panitumumab on liver lesions was blunted in patient with the *KRAS* mutation, suggesting the interplay between drug pharmacology and tumor biology. More investigations are warranted to validate this perspective.

The liver is the most common metastatic site for CRC due to its unique location in proximity to the colorectal system. Approximately 15% of CRC patients have synchronous hepatic metastases at the time of diagnosis, and another 50% will develop liver metastases in the process of treatment (39). There is a clear survival benefit in first performing a surgical resection of liver metastases when feasible, compared to resecting primary tumors first (40). One rationale behind the liver-first strategy is that liver metastasis is the main lethal factor in CRC patients. Extensive tumor hypoxia has been observed in liver metastases of CRC (41), and the hypoxic status of solid tumors is related to poor prognosis (42). The five-year survival rate is only 6% for patients with liver metastases. The early resection or reduction of liver lesions mitigate the risks of their growing beyond respectability (43). Our results were in line with the liver-first strategy and indicated the relatively higher survival benefit of favorable responses in liver metastases than in those of other organs, providing an incentive for liver-first management. For those who were not qualified for liver lesion resections, targeted therapies, such as bevacizumab or panitumumab, should be prioritized due to their favorable efficacy in liver metastases.

Our study has limitations. We analyzed data from multiple Phase III trials that had discrepancies in clinical trial design, treatment duration, and lesion evaluation frequency, resulting in high variabilities in the analyzed metrics that challenged the statistical robustness. These trials were conducted between 2002 and 2013, which made our analyses fail to reveal the current better surgical, medical, and supportive care for cancer patients. In addition, our data did not include cetuximab, another broadly prescribed targeted therapy for mCRC. Cetuximab can elicit immunologic antitumor effects, such as antibody-dependent cellular cytotoxicity, thus differing from panitumumab (44). The favorable efficacy in liver metastases by panitumumab may not be the case by cetuximab. Thus, our findings related to targeted therapy should be interpreted within the two included antibodies. Moreover, our study only analyzed the target lesions and did not include non-target lesions and new lesions, which are also critical for assessing treatment response and patient survival (9).

In conclusion, we identified the high spatiotemporal response heterogeneity across metastatic lesions in mCRC patients. The patients with higher ILH tended to have worse survival. Targeted therapies, in combination with the standard chemotherapies, showed favorable efficacy in liver metastases than chemotherapy alone, which was blunted for patients with *KRAS* mutation. Such favorable responses in the liver are more closely associated with patient long-term survival than metastatic lesions in other anatomical sites. These findings strongly support the importance of evaluating lesion-level responses for the improved assessment of drug efficacy and patient survival.

## Methods

### Data Sources

The data was extracted from the Project Sphere Database using the following criteria: Stage III or IV mCRC patients; individual lesion longitudinally measured; and recorded patient OS. In total, five Phase III trials encompassing 2,802 patients with 11,404 metastatic lesions were selected. Only 28 patients in the FOLFOX arm did not have PFS information and were excluded from the PFS analysis. The clinical trial information is summarized in Supplementary Table S1.

### Patient demographics and clinical characteristics

Patient demographic information including age, gender, race, body surface area (BSA), lactate dehydrogenase (LDH), ECOG performance score, *KRAS* status, primary tumor type, metastatic organs and number, prior medical history, RECIST response, and treatment types are listed in Table 1. The RECIST criteria-defined target lesions were also included in the analysis. Lesion anatomical sites with total lesion numbers below 50 were pooled into an “other” group including adrenal glands, chest, kidney, pancreas, peritoneum, skin, spleen, genitourinary/reproductive, and others (not specified).

### Lesion response dynamics

We developed an algorithm in MATLAB R2017b to characterize the response dynamics and the duration of the response, stable, and relapse phases at a lesion-specific level (Supplementary Fig. S1, Supplementary Codes). The criteria for defining the response, stable, and relapse phases were selected according to the RECIST v1.1. Specifically, the duration from the tumor baseline until the tumor size decreased 30% was defined as the response phase. The period, from which the lesion regrew more than 120% until the last sampling point, was described as the relapse phase. The period of neither response nor relapse was defined as the stable phase.

### Gower distance

The lesion-specific response patterns were described by five variables: three binary variables representing the presence of the response, stable, or relapse phase, and two continuous variables indicating the tumor nadir ratio and last ratio (tumor nadir size and last size before the end of trial normalized by the tumor baseline). The Gower distance of these five variables was computed as a dissimilarity measure of the response patterns across the metastases within each patient. For comparing the response patterns for lesion *i* and lesion *j*, the formula of Gower distance is

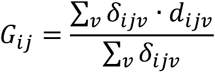

Where *v* is a variable describing the lesion response patterns, including the response, stable, relapse phase as well as the tumor nadir and last ratio. *δ*_*ijv*_ is a binary indicator equal to one when both lesions are not missing for variable *v*, otherwise equal to zero. As all the lesions in our dataset were not missing any variables, ∑_*v*_ *δ*_*ijv*_ is the number of variables and equal to five.

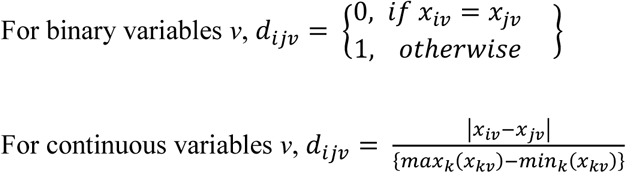

Where the equation *max*_*k*_(*x*_*kv*_) − *min*_*k*_(*x*_*kv*_) represents the range of the variable *v*. The *d*_*ijv*_ is set to zero if *max*_*k*_(*x*_*kv*_) − *min*_*k*_(*x*_*kv*_) = 0.

Lesion-level heterogeneity was quantified by the Gower distances across the metastases. The average Gower distances of all the metastases within one patient represents the patient-level heterogeneity.

### Statistical analysis

Comparisons of two continuous variables were performed using the Mann-Whitney test. For multiple comparisons, we performed the Kruskal-Wallis test, followed by Dunn’s tests with Bonferroni corrections. PFS (defined as the start of therapies until RECIST-defined progression or death) and OS (defined as the start of therapies until patient death) among the groups were depicted using Kaplan-Meier curves and compared using log-rank tests. The estimated hazard ratio (HR) and p value of the Gower distances – controlling for other confounding factors – were calculated in the Cox proportional hazard model stratified by treatment type. The mean (standard deviation) and median (interquartile range) of each group compared are presented in Supplementary Tables S2-S12, and a p value < 0.05 with a two-sided alternative was considered statistically significant. All the statistical tests were performed using R 3.5.1 or GraphPad Prism 8. The figures were made in R 3.5.1, GraphPad Prism 8 and BioRender software.

## Data Availability

All data has made available in the supplementary materials

## Abbreviations

RECIST: Response Evaluation Criteria in Solid Tumors
LN: lymph nodes
GI: gastrointestinal
CRC/mCRC: colorectal cancer/metastatic colorectal cancer
PFS: progression-free survival
OS: overall survival
ILH: inter-lesion heterogeneity
Beva: bevacizumab
Pani: panitumumab
Chemo: chemotherapy
FOLFOX: folinic acid (leucovorin), fluorouracil (5-FU), and oxaliplatin (Eloxatin)
FOLFIRI: folinic acid (leucovorin), fluorouracil (5-FU), and irinotecan (Camptosar)
MAPK: mitogen-activated protein kinase
EGFR: epidermal growth factor receptor
VEGF: vascular endothelial growth factor
BSA: body surface area
LDH: lactate dehydrogenase
HR: hazard ratio

