## Supplementary Material for "Spatiotemporal Response Heterogeneity Across Metastatic Lesions Informs Drug Efficacy and Patient Survival in Colorectal Cancer"

**Supplementary Figures**

**Supplementary Figure S1**. Conceptual workflow of the lesion pattern definition algorithm.

**Supplementary Figure S2**. The response patterns of metastatic lesions in patients with high or low Gower distance.

**Supplementary Figure S3**. Treatment efficacy in Beva + Chemo versus Chemo only.

**Supplementary Figure S4**. Treatment efficacy in Pani + Chemo versus Chemo only.

**Supplementary Figure S5**. Treatment efficacy in patients with *KRAS* mutations versus wild-type.

**Supplementary Figure S6**. The LN or lung metastases responses were not associated with patient survival.

**Supplementary Tables**

**Supplementary Table S1**. Data Source.

**Supplementary Table S2-12**. The mean (standard deviation) and median (interquartile range) of the data analyzed in the figures.

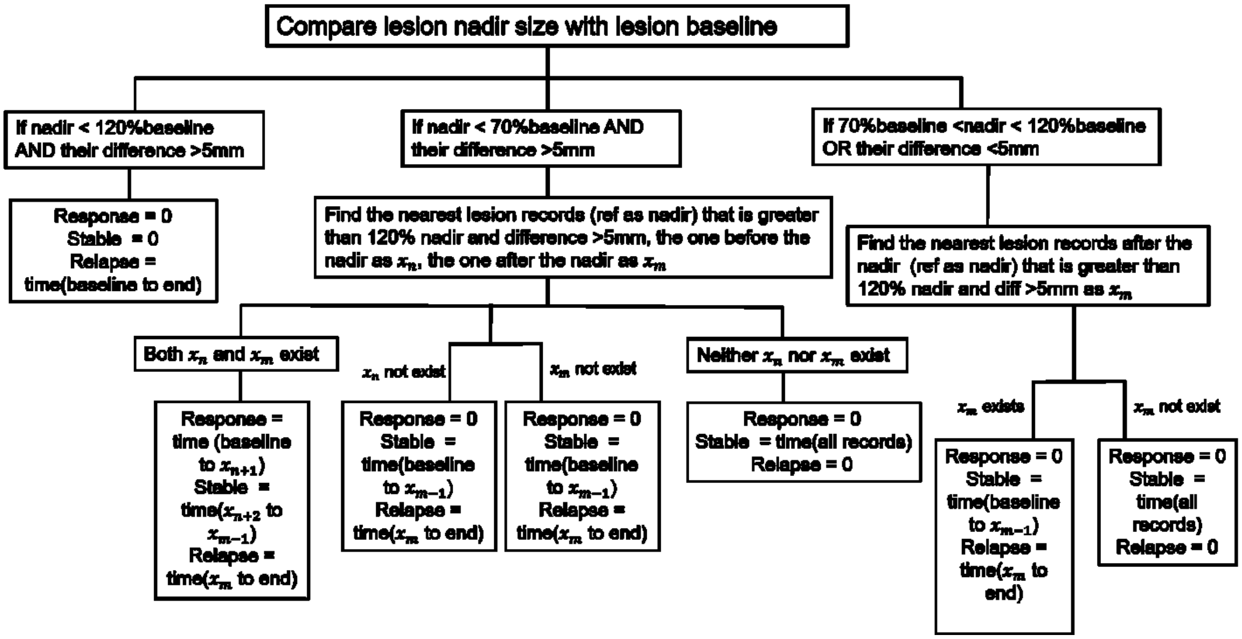


**Supplementary Figure S1**. **Conceptual workflow of the lesion pattern characterization algorithm.** We developed an algorithm in MATLAB to characterize lesion response patterns and measure the durations of response, stable and relapse phase.

(A)

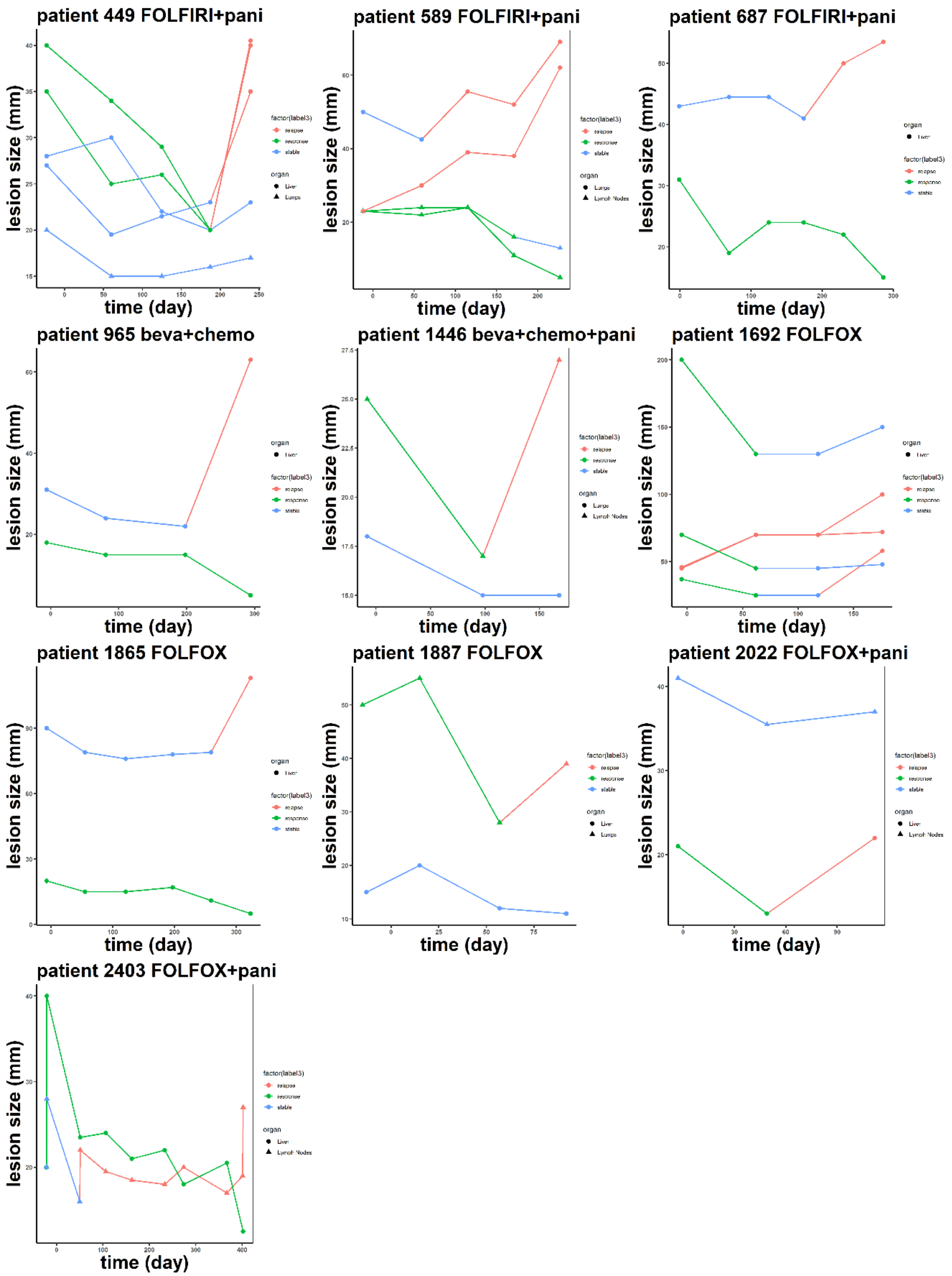
patient 449 FOLFIRI+pani
patient 589 FOLFIRI+pani
patient 687 FOLFIRI+pani
patient 965 beva+chemo
patient 1446 beva+chemo+pani
patient 1692 FOLFOX
patient 1865 FOLFOX
patient 1887 FOLFOX
patient 2022 FOLFOX+pani
patient 2403 FOLFOX+pani
lesion size (mm)
time (day)
factor(label3)
organ

(B)

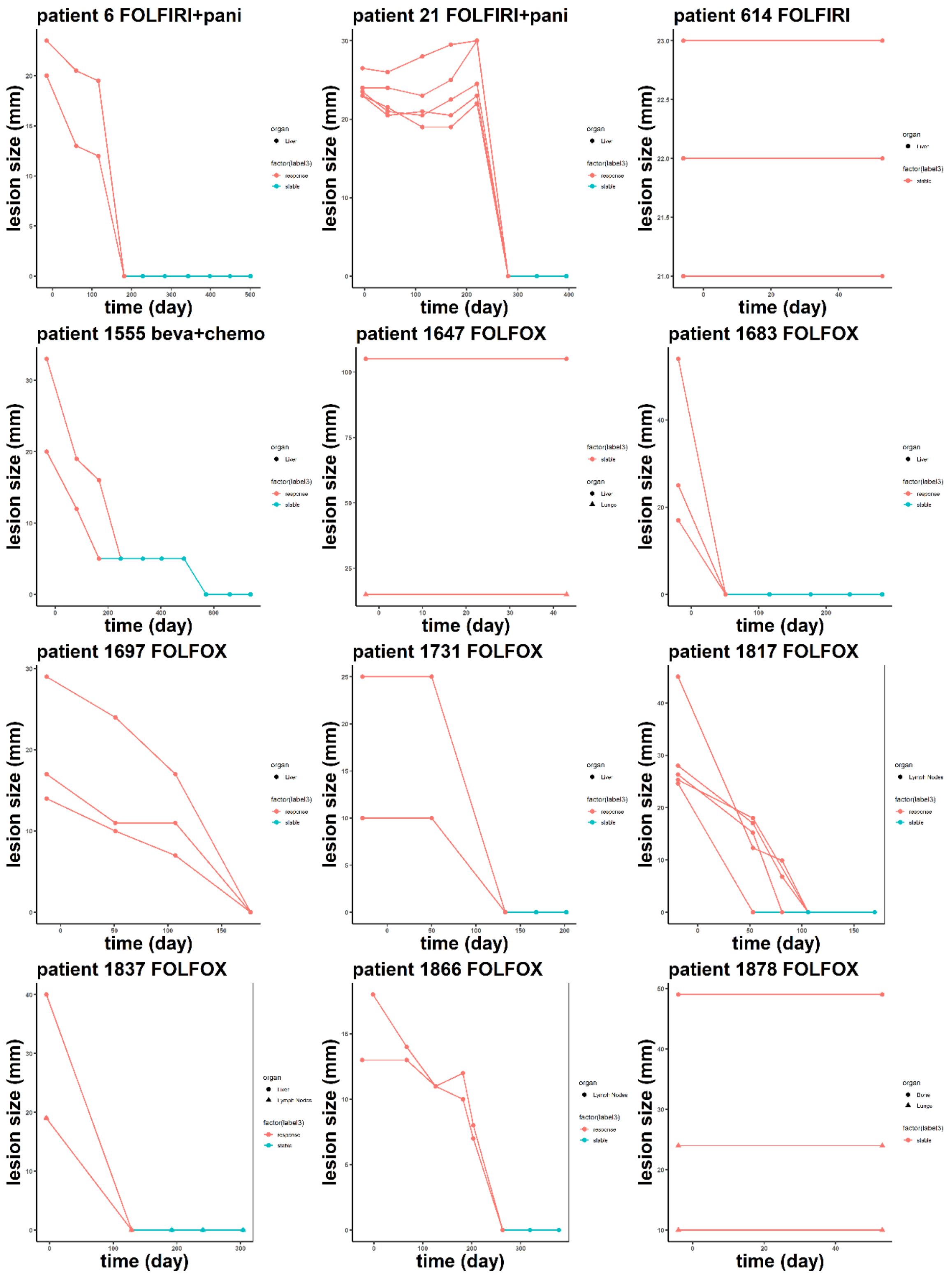
patient 6 FOLFIRI+pani
patient 21 FOLFIRI+pani
patient 614 FOLFIRI
patient 1555 beva+chemo
patient 1647 FOLFOX
patient 1683 FOLFOX
patient 1697 FOLFOX
patient 1731 FOLFOX
patient 1817 FOLFOX
patient 1837 FOLFOX
patient 1866 FOLFOX
patient 1878 FOLFOX
lesion size (mm)
time (day)

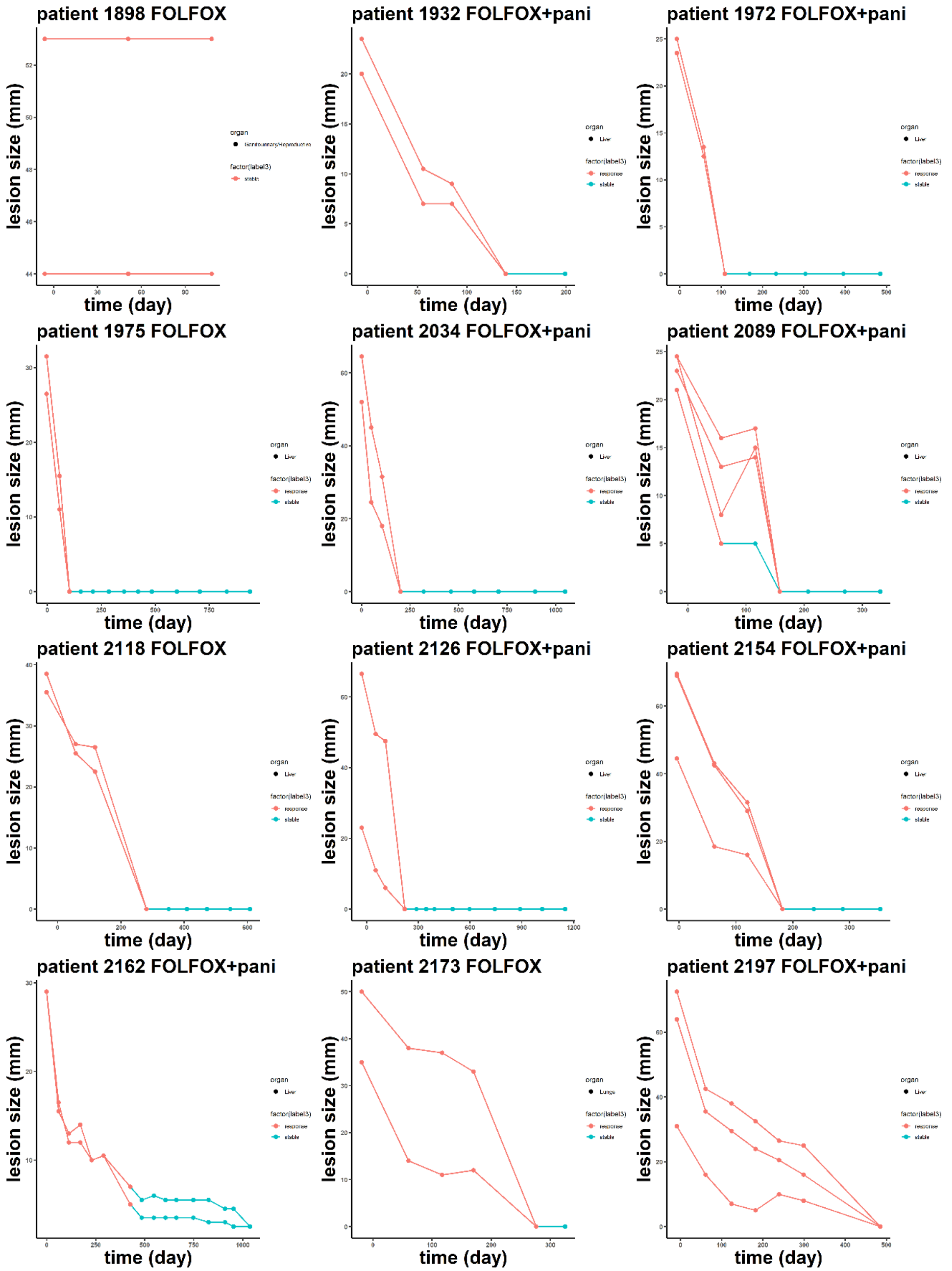
patient 1898 FOLFOX
patient 1932 FOLFOX+pani
patient 1972 FOLFOX+pani
patient 1975 FOLFOX
patient 2034 FOLFOX+pani
patient 2089 FOLFOX+pani
patient 2118 FOLFOX
patient 2126 FOLFOX+pani
patient 2154 FOLFOX+pani
patient 2162 FOLFOX+pani
patient 2173 FOLFOX
patient 2197 FOLFOX+pani
lesion size (mm)
time (day)
organ
factor(label3)

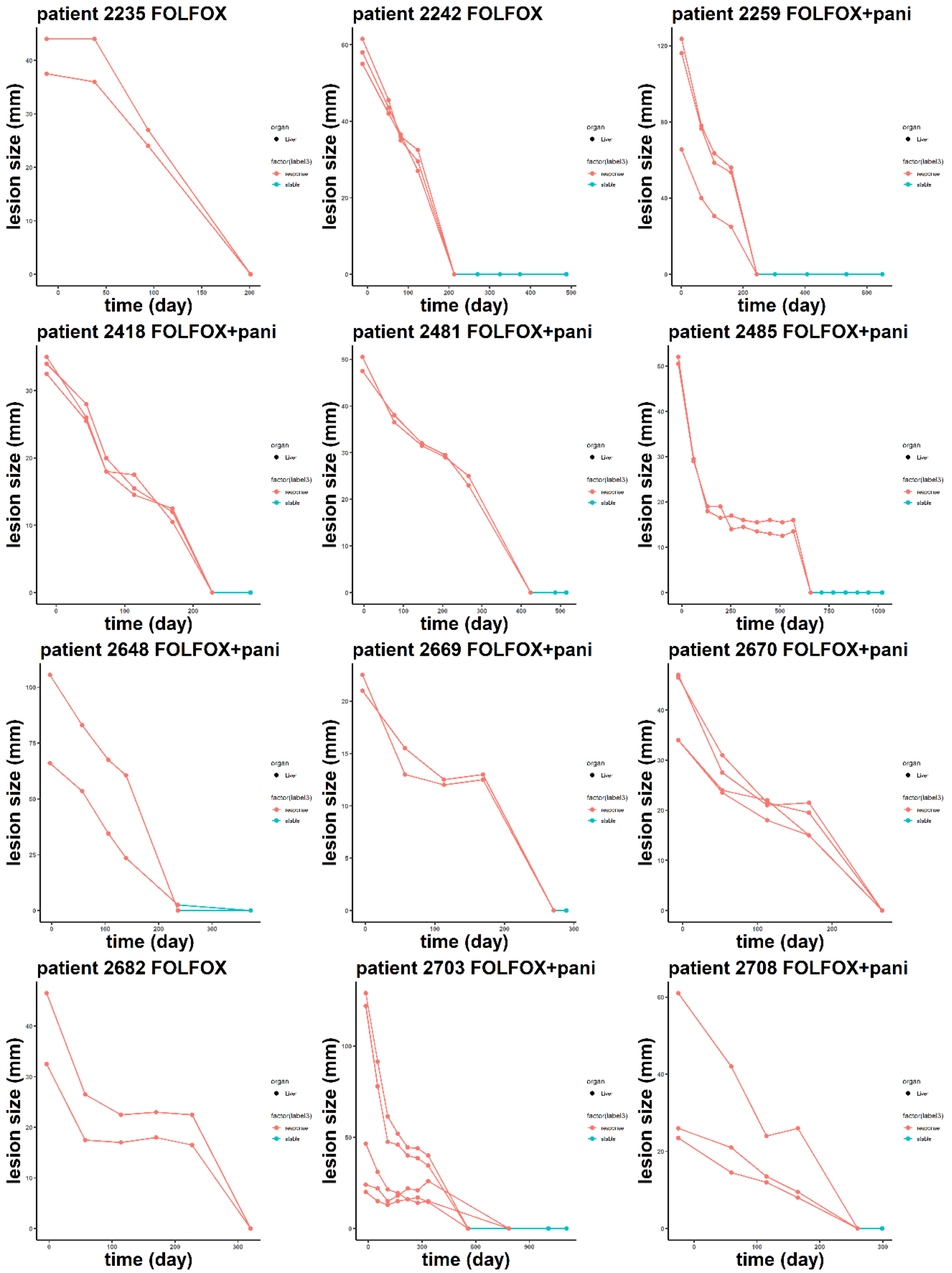
patient 2235 FOLFOX
patient 2242 FOLFOX
patient 2259 FOLFOX+pani
patient 2418 FOLFOX+pani
patient 2481 FOLFOX+pani
patient 2485 FOLFOX+pani
patient 2648 FOLFOX+pani
patient 2669 FOLFOX+pani
patient 2670 FOLFOX+pani
patient 2682 FOLFOX
patient 2703 FOLFOX+pani
patient 2708 FOLFOX+pani
lesion size (mm)
time (day)
organ
factor(label3)

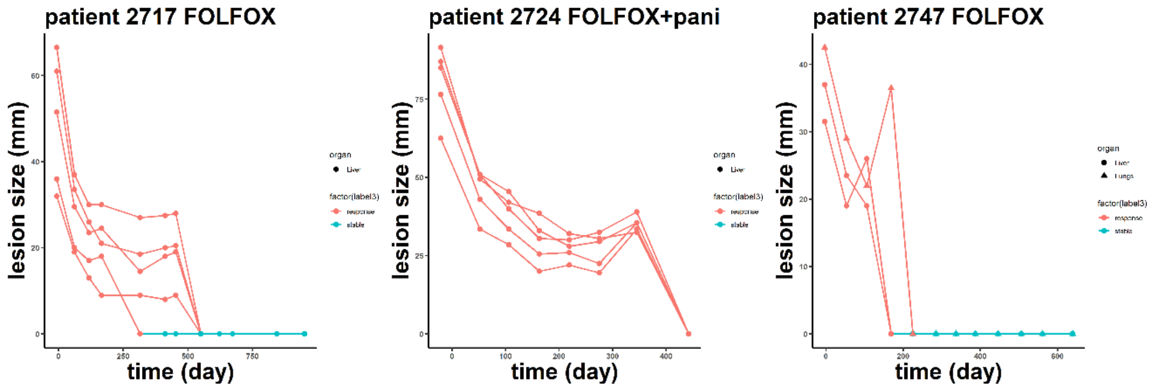


**Supplementary Figure S2**. **The response patterns of metastatic lesions in patients with high Gower distance (A) or low Gower distance (B).** The Gower distances in (A) range from 0.45 to 0.5 (top 10 in our data set) and the Gower distances in (B) all equal to 0. Patient identification number and corresponding treatment were labeled in each subpanel. Lesions in different anatomical sites were labeled in different symbols. The response, stable, and relapse patterns were plotted in red, green, and blue respectively.

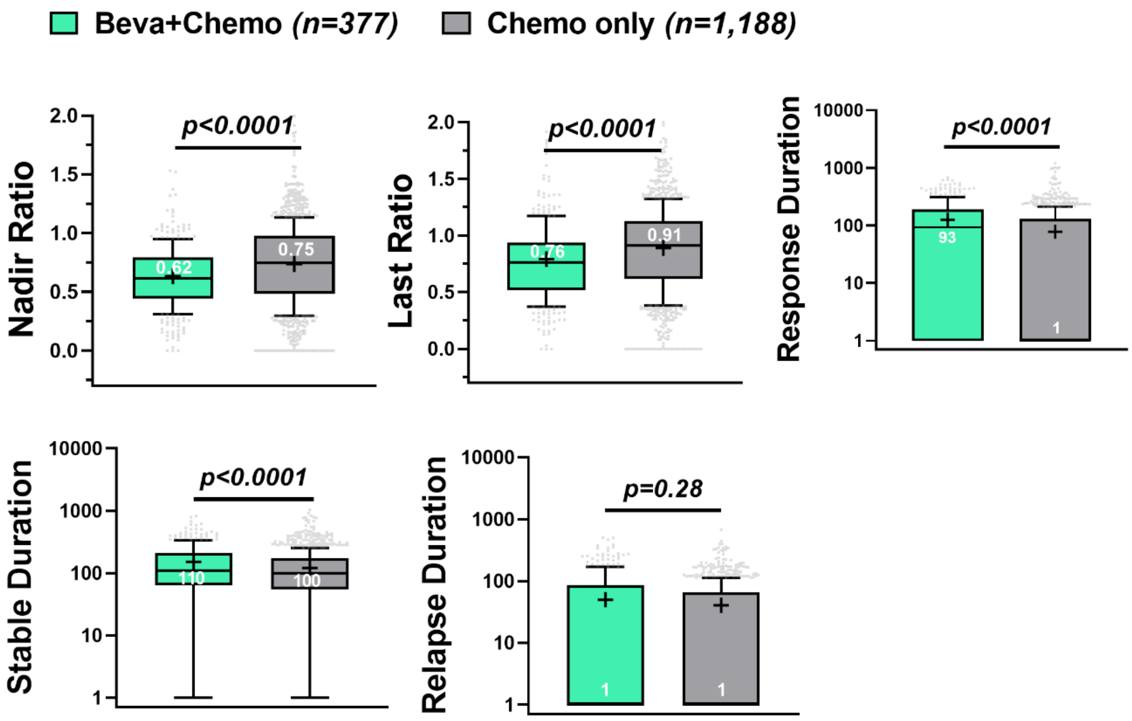


**Supplementary Figure S3**. **Treatment efficacy in Beva + Chemo versus Chemo only.** The nadir ratio, last ratio, response duration, stable duration and relapse duration of the total tumor burden at patient-level in the bevacizumab arm (Beva + Chemo) versus chemotherapy only arm (Chemo only).

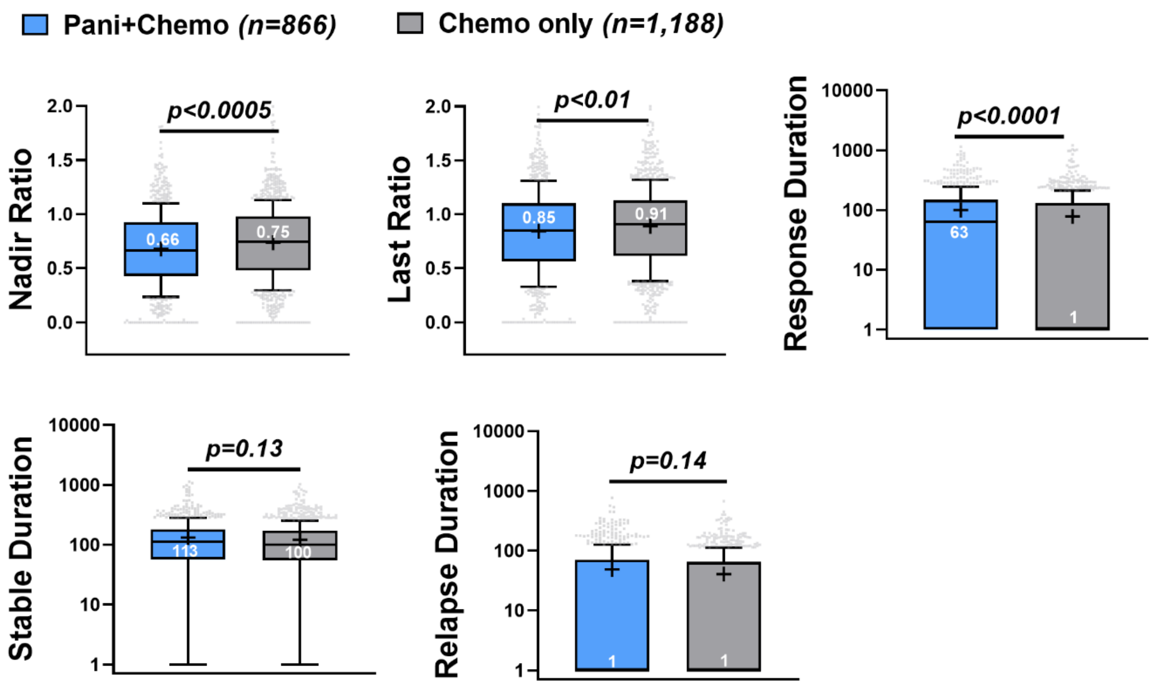


**Supplementary Figure S4**. **Treatment efficacy in Pani + Chemo versus Chemo only.** The nadir ratio, last ratio, response duration, stable duration and relapse duration of the total tumor burden at patient-level in the panitumumab arm (Pani + Chemo) versus chemotherapy only arm (Chemo only).

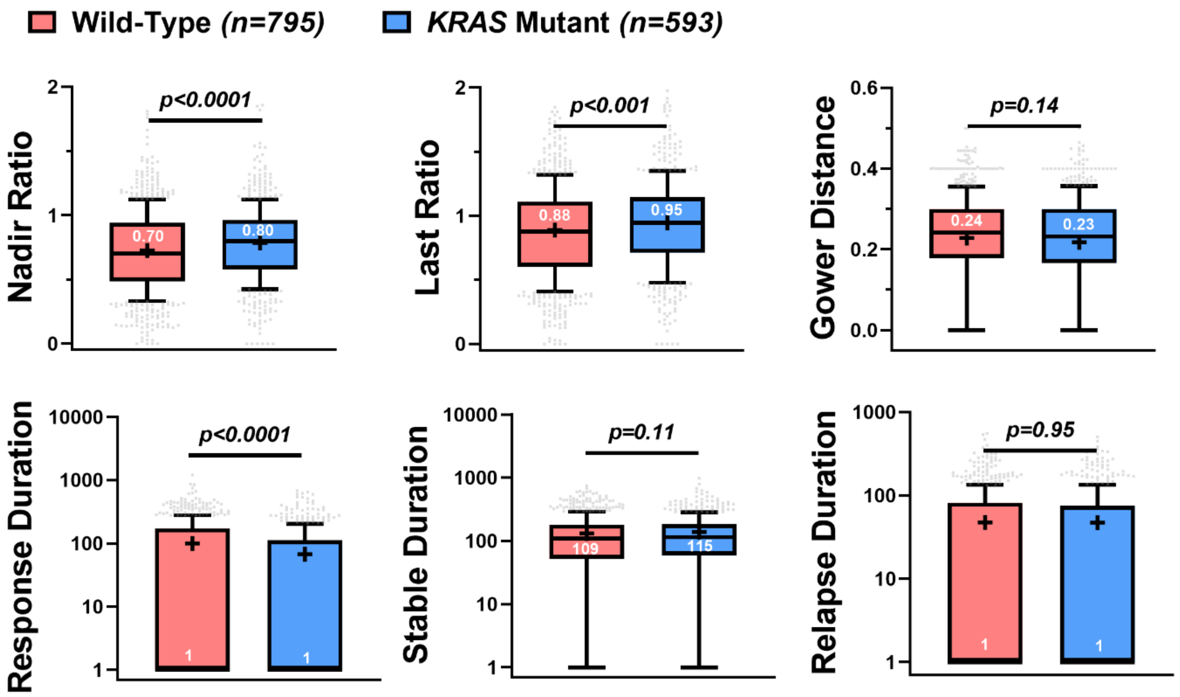


**Supplementary Figure S5**. **Treatment efficacy in the patients with *KRAS* mutations versus wild-type.** The nadir ratio, last ratio, Gower distance, response duration, stable duration and relapse duration of the total tumor burden in the wild-type patients (n = 795) vs *KRAS* mutant patients (n = 593).

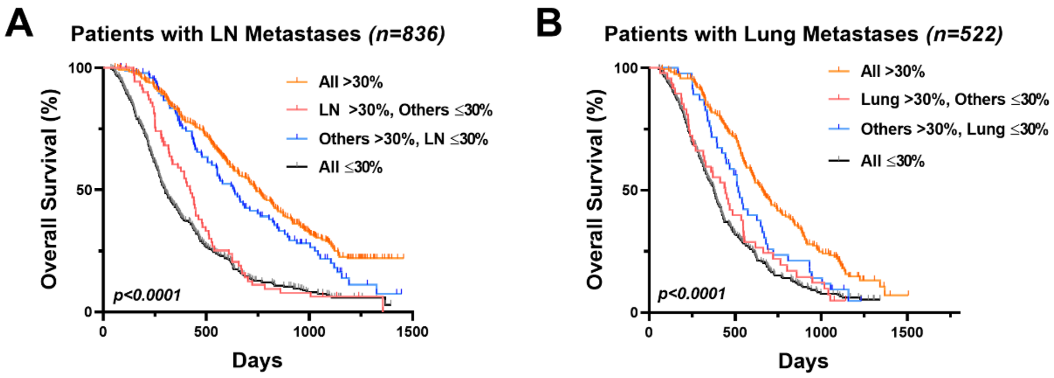


**Supplementary Figure S6**. **The LN or lung metastases responses were not associated with patient survival.** Kaplan-Meier curves of the OS in patients with LN metastases (A) or lung metastases (B). The patients were divided into four subgroups in (A): all lesions shrinkage greater than 30% threshold (all > 30%, n = 334); LN lesions shrinkage > 30% while other lesions shrinkage ≤ 30% (LN > 30%, others ≤ 30%; n = 72); LN lesions shrinkage ≤ 30% while other lesions shrinkage > 30% (others >30%, LN ≤ 30%; n = 100); all lesions shrinkage ≤ 30% (all ≤ 30%, n = 330). The patients in (B) were divided in similar subgroups: all > 30% (n = 190); lung > 30%, others ≤ 30% (n = 49); others >30%, lung ≤ 30% (n = 46); all ≤ 30% (n = 237).

**Supplementary Data Tables**.

**Supplementary Table S1**. **Data Source.** Descriptive information about the five randomized clinical trials in metastatic colorectal cancer collected from Project Data Sphere.

| | **Experimental agent being tested** | **Control treatment** | **Patients included (No.)** | **Masking Method** | **Study phase** | **Study Start date** | **Study end date** | **ClinicalTrial.gov identifier** |
|---|---|---|---|---|---|---|---|---|
| Amgen 2006 263 | Panitumumab plus FOLFIRI | FOLFIRI | 844 | Open Label | 3 | June 2006 | November 2010 | NCT00339183 |
| Amgen 2005 262 (PACCE) | Bevacizumab plus Panitumumab plus chemotherapy | Bevacizumab plus chemotherapy | 748 | Open Label | 3 | June 2005 | May 2009 | NCT00115765 |
| Amgen 2006 264 (PRIME) | FOLFOX plus Panitumumab | FOLFOX | 884 | Open Label | 3 | August 2006 | March 2013 | NCT00364013 |
| SanofiU 2002 137 (XENOX) | FOLFOX plus Xaliproden | FOLFOX | 130 | Double blinded | 3 | July 2002 | May 2004 | NCT00272051 |
| SanofiU 2005 136 | FOLFOX plus Xaliproden | FOLFOX | 196 | Double blinded | 3 | December 2005 | October 2009 | NCT00305188 |

**Table S2**. The mean (standard deviation) and the median (interquartile range) of the Gower distances at lesion-level across anatomical sites.

| Organ (No.) | Mean (sd) | Median (IQR) | Organ (No.) | Mean (sd) | Median (IQR) |
|---|---|---|---|---|---|
| Abdomen (399) | 0.27 (0.12) | 0.26 (0.14) | Bone (166) | 0.27 (0.13) | 0.27 (0.15) |
| GI (130) | 0.28 (0.14) | 0.28 (0.18) | Liver (7,449) | 0.25 (0.12) | 0.23 (0.14) |
| Lungs (1,169) | 0.25 (0.11) | 0.23 (0.14) | Lymph Nodes (1,833) | 0.26 (0.12) | 0.24 (0.15) |
| Other (258) | 0.25 (0.13) | 0.23 (0.15) | | | |

**Table S3**. The mean (standard deviation) and the median (interquartile range) of the Gower distances at patient-level across treatment types.

| Treatment (No.) | Mean (sd) | Median (IQR) | Treatment (No.) | Mean (sd) | Median (IQR) |
|---|---|---|---|---|---|
| Targeted+Chemo (1,614) | 0.22 (0.12) | 0.23 (0.13) | Chemo (1,188) | 0.23 (0.12) | 0.24 (0.13) |
| Beva+Chemo (377) | 0.22 (0.12) | 0.23 (0.13) | Beva+Chemo+Pani (371) | 0.22 (0.12) | 0.24 (0.13) |
| FOLFIRI (419) | 0.22 (0.11) | 0.23 (0.13) | FOLFIRI+Pani (425) | 0.23 (0.12) | 0.24 (0.13) |
| FOLFOX (769) | 0.23 (0.12) | 0.25 (0.13) | FOLFOX+Pani (441) | 0.21 (0.12) | 0.22 (0.13) |

**Table S4.** The mean (standard deviation) and the median (interquartile range) of the response pattern parameters at patient-level in the bevacizumab arm (Beva+Chemo) vs chemotherapy arm (Chemo only). The durations were added with one to avoid zero values in logarithmic plots in all the tables below.

| | Beva+Chemo (n = 377) | | Chemo only (n = 1,188) | |
|---|---|---|---|---|
| | Mean (sd) | Median (IQR) | Mean (sd) | Median (IQR) |
| Nadir Ratio | 0.63 (0.33) | 0.62 (0.35) | 0.74 (0.36) | 0.75 (0.50) |
| Last Ratio | 0.79 (0.53) | 0.76 (0.42) | 0.89 (0.42) | 0.91 (0.48) |
| Gower distance | 0.22 (0.12) | 0.23 (0.13) | 0.23 (0.12) | 0.24 (0.13) |
| Response duration | 126 (141) | 93 (188) | 78 (125) | 1 (129) |
| Stable duration | 151 (137) | 110 (146) | 121 (119) | 100 (117) |
| Relapse duration | 50 (88) | 1 (86) | 41 (61) | 1 (64) |

**Table S5.** The mean (standard deviation) and the median (interquartile range) of the lesion-level nadir ratios in the bevacizumab arm (Beva+Chemo) vs chemotherapy arm (Chemo only).

| Beva+Chemo (n = 1,634) | | | Chemo only (n = 4,764) | | |
|---|---|---|---|---|---|
| Organ (No.) | Mean (sd) | Median (IQR) | Organ (No.) | Mean (sd) | Median (IQR) |
| Abdomen (63) | 0.58 (0.28) | 0.56 (0.40) | Abdomen (170) | 0.89 (0.33) | 0.92 (0.40) |
| Bone (8) | 0.62 (0.27) | 0.63 (0.42) | Bone (91) | 0.85 (0.33) | 0.88 (0.37) |
| GI (10) | 0.71 (0.28) | 0.72 (0.44) | GI (65) | 0.76 (0.29) | 0.81 (0.46) |
| Liver (1,031) | 0.60 (0.29) | 0.58 (0.39) | Liver (3,128) | 0.72 (0.39) | 0.71 (0.57) |
| Lungs (146) | 0.71 (0.23) | 0.73 (0.28) | Lungs (513) | 0.73 (0.31) | 0.79 (0.44) |
| Lymph Nodes (353) | 0.64 (0.23) | 0.66 (0.38) | Lymph Nodes (646) | 0.71 (0.33) | 0.74 (0.45) |
| Other (23) | 0.72 (0.26) | 0.69 (0.48) | Other (151) | 0.81 (0.36) | 0.86 (0.40) |

**Table S6.** The mean (standard deviation) and the median (interquartile range) of the lesion-level Gower distances in the bevacizumab arm (Beva+Chemo) vs chemotherapy arm (Chemo only).

| Beva+Chemo (n = 1,634) | | | Chemo only (n = 4,764) | | |
|---|---|---|---|---|---|
| Organ (No.) | Mean (sd) | Median (IQR) | Organ (No.) | Mean (sd) | Median (IQR) |
| Abdomen (63) | 0.26 (0.13) | 0.29 (0.11) | Abdomen (170) | 0.26 (0.12) | 0.25 (0.12) |
| Bone (8) | 0.33 (0.10) | 0.36 (0.16) | Bone (91) | 0.28 (0.13) | 0.28 (0.16) |
| GI (10) | 0.26 (0.13) | 0.29 (0.14) | GI (65) | 0.27 (0.14) | 0.27 (0.18) |
| Liver (1,031) | 0.24 (0.11) | 0.22 (0.13) | Liver (3,128) | 0.25 (0.12) | 0.24 (0.14) |
| Lungs (146) | 0.25 (0.12) | 0.24 (0.16) | Lungs (513) | 0.24 (0.12) | 0.22 (0.14) |
| Lymph Nodes (353) | 0.26 (0.11) | 0.25 (0.15) | Lymph Nodes (646) | 0.26 (0.13) | 0.24 (0.16) |
| Other (23) | 0.22 (0.11) | 0.23 (0.13) | Other (151) | 0.25 (0.13) | 0.22 (0.16) |

**Table S7.** The mean (standard deviation) and the median (interquartile range) of the lesion-level response pattern parameters in liver, lung and LN metastases in the bevacizumab arm (Beva+Chemo).

| | Liver (n = 1,031) | | Lungs (n = 146) | | Lymph Nodes (n = 353) | |
|---|---|---|---|---|---|---|
| | Mean (sd) | Medina (IQR) | Mean (sd) | Medina (IQR) | Mean (sd) | Medina (IQR) |
| Nadir Ratio | 0.60 (0.29) | 0.58 (0.39) | 0.71 (0.23) | 0.73 (0.28) | 0.64 (0.23) | 0.66 (0.38) |
| Last Ratio | 0.76 (0.44) | 0.72 (0.48) | 0.89 (0.36) | 0.85 (0.39) | 0.74 (0.28) | 0.75 (0.37) |
| Gower distance | 0.24 (0.11) | 0.22 (0.13) | 0.25 (0.12) | 0.24 (0.16) | 0.26 (0.11) | 0.25 (0.15) |

| | | | | | | |
|---|---|---|---|---|---|---|
| Response duration | 109 (116) | 92 (175) | 54 (87) | 1 (91) | 78 (96) | 78 (113) |
| Stable duration | 173 (132) | 159 (154) | 216 (139) | 186 (235) | 223 (161) | 185 (226) |
| Relapse duration | 37 (71) | 1 (54) | 33 (79) | 1 (0) | 25 (69) | 1 (0) |

**Table S8.** The mean (standard deviation) and the median (interquartile range) of the lesion-level response pattern parameters in liver, lung and LN metastases in the chemotherapy arm (Chemo Only).

| | Liver (n = 3,128) | | Lungs (n = 513) | | Lymph Nodes (n = 646) | |
|---|---|---|---|---|---|---|
| | Mean (sd) | Medina (IQR) | Mean (sd) | Medina (IQR) | Mean (sd) | Medina (IQR) |
| Nadir Ratio | 0.72 (0.39) | 0.71 (0.57) | 0.73 (0.31) | 0.79 (0.44) | 0.71 (0.33) | 0.74 (0.45) |
| Last Ratio | 0.88 (0.44) | 0.89 (0.63) | 0.87 (0.36) | 0.91 (0.44) | 0.81 (0.38) | 0.84 (0.43) |
| Gower distance | 0.25 (0.12) | 0.24 (0.14) | 0.24 (0.12) | 0.22 (0.14) | 0.26 (0.13) | 0.24 (0.16) |
| Response duration | 74 (105) | 1 (124) | 45 (72) | 1 (72) | 62 (113) | 1 (79) |
| Stable duration | 116 (119) | 92 (109) | 142 (105) | 122 (122) | 153 (146) | 118 (128) |
| Relapse duration | 39 (58) | 1 (63) | 23 (42) | 1 (52) | 19 (44) | 1 (0) |

**Table S9.** The mean (standard deviation) and the median (interquartile range) of the response pattern parameters at patient-level in the panitumumab arm (Pani+Chemo) vs chemotherapy arm (Chemo only).

| | Pani+Chemo (n = 866) | | Chemo only (n = 1,188) | |
|---|---|---|---|---|
| | Mean (sd) | Median (IQR) | Mean (sd) | Median (IQR) |
| Nadir Ratio | 0.68 (0.36) | 0.66 (0.49) | 0.74 (0.36) | 0.75 (0.50) |
| Last Ratio | 0.84 (0.44) | 0.85 (0.54) | 0.89 (0.42) | 0.91 (0.48) |
| Gower distance | 0.22 (0.12) | 0.23 (0.13) | 0.23 (0.12) | 0.24 (0.13) |
| Response duration | 100 (139) | 63 (147) | 78 (125) | 1 (129) |
| Stable duration | 132 (135) | 113 (123) | 121 (119) | 100 (117) |
| Relapse duration | 49 (77) | 1 (69) | 41 (61) | 1 (64) |

**Table S10.** The mean (standard deviation) and the median (interquartile range) of the lesion-level nadir ratios across anatomical sites in the panitumumab arm (Pani + Chemo) vs chemotherapy arm (Chemo only).

| Pani+Chemo (n = 3,500) | | | Chemo only (n = 4,764) | | |
|---|---|---|---|---|---|
| Organ (No.) | Mean (sd) | Median (IQR) | Organ (No.) | Mean (sd) | Median (IQR) |
| Abdomen (116) | 0.83 (0.35) | 0.89 (0.43) | Abdomen (170) | 0.89 (0.33) | 0.92 (0.40) |
| Bone (55) | 0.83 (0.28) | 0.82 (0.50) | Bone (91) | 0.85 (0.33) | 0.88 (0.37) |
| GI (44) | 0.66 (0.34) | 0.70 (0.58) | GI (65) | 0.76 (0.29) | 0.81 (0.46) |
| Liver (2,422) | 0.64 (0.37) | 0.60 (0.52) | Liver (3,128) | 0.72 (0.39) | 0.71 (0.57) |
| Lungs (266) | 0.72 (0.24) | 0.69 (0.37) | Lungs (513) | 0.73 (0.31) | 0.79 (0.44) |
| Lymph Nodes (534) | 0.68 (0.29) | 0.70 (0.49) | Lymph Nodes (646) | 0.71 (0.33) | 0.74 (0.45) |
| Other (63) | 0.92 (0.31) | 0.95 (0.36) | Other (151) | 0.81 (0.36) | 0.86 (0.40) |

**Table S11.** The mean (standard deviation) and the median (interquartile range) of the lesion-level Gower distances across anatomical sites in the panitumumab arm (Pani + Chemo) vs chemotherapy arm (Chemo only).

| Pani+Chemo (n = 3,500) | | | Chemo only (n = 4,764) | | |
|---|---|---|---|---|---|
| Organ (No.) | Mean (sd) | Median (IQR) | Organ (No.) | Mean (sd) | Median (IQR) |
| Abdomen (116) | 0.27 (0.12) | 0.26 (0.14) | Abdomen (170) | 0.26 (0.12) | 0.25 (0.12) |
| Bone (55) | 0.27 (0.11) | 0.27 (0.13) | Bone (91) | 0.28 (0.13) | 0.28 (0.16) |
| GI (44) | 0.29 (0.15) | 0.32 (0.20) | GI (65) | 0.27 (0.14) | 0.27 (0.18) |
| Liver (2,422) | 0.24 (0.11) | 0.22 (0.14) | Liver (3,128) | 0.25 (0.12) | 0.24 (0.14) |
| Lungs (266) | 0.25 (0.10) | 0.24 (0.12) | Lungs (513) | 0.24 (0.12) | 0.22 (0.14) |
| Lymph Nodes (534) | 0.27 (0.12) | 0.25 (0.15) | Lymph Nodes (646) | 0.26 (0.13) | 0.24 (0.16) |
| Other (63) | 0.26 (0.12) | 0.23 (0.14) | Other (151) | 0.25 (0.13) | 0.22 (0.16) |

**Table S12.** The mean (standard deviation) and the median (interquartile range) of the lesion-level response pattern parameters in liver, lung and LN metastases in the panitumumab arm (Pani + Chemo).

| | Liver (n = 2,422) | | Lungs (n = 266) | | Lymph Nodes (n = 534) | |
|---|---|---|---|---|---|---|
| | Mean (sd) | Median (IQR) | Mean (sd) | Median (IQR) | Mean (sd) | Median (IQR) |
| Nadir over baseline | 0.64 (0.37) | 0.60 (0.52) | 0.72 (0.24) | 0.69 (0.37) | 0.68 (0.29) | 0.70 (0.49) |
| Last over baseline | 0.82 (0.45) | 0.80 (0.61) | 0.91 (0.33) | 0.90 (0.41) | 0.80 (0.34) | 0.81 (0.46) |
| Gower distance | 0.24 (0.11) | 0.22 (0.14) | 0.25 (0.10) | 0.24 (0.12) | 0.27 (0.12) | 0.25 (0.15) |
| Response duration | 103 (130) | 68 (146) | 52 (70) | 53 (77) | 65 (92) | 46 (112) |

| | | | | | | |
|---|---|---|---|---|---|---|
| Stable duration | 136 (142) | 113 (123) | 152 (99) | 127 (125) | 156 (135) | 119 (135) |
| Relapse duration | 43 (69) | 1 (63) | 42 (77) | 1 (58) | 32 (70) | 1 (49) |
